# Hypoxic Microenvironment Promotes PTBP1 Lactonization and IGF2BP2 Read Defects Mediate the Development of Preeclampsia

**DOI:** 10.1101/2023.07.05.23292275

**Authors:** Hongmei Qu, Xiaoyan Li, Qian Li, Xiaoming Yang, Yan Feng, Li Yu, Liping Qu, Linsong Mu, Yanfen Zou, Yongli Chu

**Affiliations:** Departments of Obstetrics and Gynecology, The Affiliated Yantai Yuhuangding Hospital of Qingdao University, Yantai, Shandong 264000, P.R. China; Departments of Scientific Research, The Affiliated Yantai Yuhuangding Hospital of Qingdao University, Yantai, Shandong 264000, P.R. China; Department of clinical nutrition, The Affiliated Yantai Yuhuangding Hospital of Qingdao University, Yantai, Shandong 264000, P.R. China; Department of Central Ward Operating Room, The Affiliated Yantai Yuhuangding Hospital of Qingdao University, Yantai, Shandong 264000, P.R. China; Department of General Surgery and Pediatric Surgery, The Affiliated Yantai Yuhuangding Hospital of Qingdao University, Yantai, Shandong 264000, P.R. China

**Author notes:** Corresponding author: Linsong Mu, Yanfen Zou, Yongli Chu. Contribute equally as co-first author (Hongmei Qu, Xiaoyan Li, Qian Li).

**Keywords:** single-cell transcriptomics, preeclampsia, chorionic trophoblast, hypoxia, oxidative stress, mitochondrial autophagy

## Abstract

**Objective:** As an idiopathic hypertensive disorder of pregnancy, pre-eclampsia (PE) remains a major cause of maternal and neonatal morbidity and mortality, with no effective strategy for causal treatment.

**Methods:** This study was performed by downloading the Gene Expression Omnibus (GEO) database (http://www.ncbi.nlm.nih.gov/geo/) based on the GSE173193 dataset, including single-cell sequencing data from placental samples of two PE patients and two normal controls. Placental cell subpopulations and their transcriptional heterogeneity were compared between PE and healthy pregnancies, and the mechanisms of PE cell dynamics in the hypoxic microenvironment were confirmed by in vitro experiments.

**Results:** In this study, we constructed a large-scale single-cell transcriptome ecological landscape of 26,416 cells from healthy pregnant and PE patients placenta and further identified a PE-specific CSNK2B-positive subpopulation of chorionic villous trophoblast (EVT) cells. Specifically, this study revealed that the EVT subpopulation PTBP1 was inactivated by lactonization in the hypoxic microenvironment, resulting in low expression of the N6-methyladenosine (m^6^A) reading protein IGF2BP2. On the basis of this, low expression of IGF2BP2 inhibits mitochondrial autophagy, causes the accumulation of damaged mitochondria, exacerbates lactic acid accumulation while inducing EVT apoptosis on the one hand. In particular, hypoxia may initially promote oxidative stress through the production of mitochondrial reactive oxygen species. on the other hand, it inhibits EVT adherent spot signaling, decreases EVT invasive ability, leads to impaired placental spiral vessel recast, and promotes PE disease process. In addition, there are interactions between abnormal metabolic signaling of PE-specific EVT subpopulations and microenvironmental immune cells, which activate metabolic inflammation.

**Conclusion:** The present study not only provides a new cell biological and genetic basis for elucidating the pathogenesis of PE, but also contributes to the design of an allopathic treatment strategy for PE.

## Introduction

Pre-eclampsia (PE) is a hypertensive disorder specific to pregnancy and a major cause of maternal and neonatal morbidity and mortality (4%-5%), manifesting as new-onset hypertension and proteinuria or end-organ damage after 20 weeks of gestation (1–3). Globally, PE affects about 2-8% of pregnancies, killing about 76,000 pregnant women and 500,000 fetuses each year, and even causing up to 20% of preterm birth events (4–6). However, as of today, PE remains incurable, with blood pressure control being the main clinical intervention and placental delivery remaining the only definitive treatment (7).

To date, the pathogenesis of PE is not fully understood, but abnormal placental development plays an important role in the disease (8). During the early stages of conception, trophoblast cells differentiate into cytotrophoblast cells (CTB) and syncytial trophoblast cells (ST), followed by the formation of extravillous trophoblast (EVT) from CTB The EVT is able to invade the endometrium and remodel the uterine spiral arteries to allow successful embryo implantation and development (9). In addition, this process involves many cells at the maternal-fetal interface, including some immune microenvironmental cells in addition to the trophoblast system described above (10), which fully reflects the heterogeneous nature of placental cells.

Numerous studies have confirmed that hypoxia is closely associated with the development of PE. decreased placental blood perfusion in the uterus of PE patients, and the expression of hypoxia-related genes showed a striking correlation with the placenta of PE patients, with consistent results in both in vivo and in vitro models of placental hypoxia (11). The finding that rodent and primate models of uteroplacental ischemia and hypoxia (12, 13) and a rat model of uteroplacental ischemia or hypoperfusion (12) showed the same clinical signs as PE further confirms the relevance of hypoxia to PE. Previous studies have demonstrated that early gestational trophoblast cells produce large amounts of lactate through aerobic glycolysis at the maternal-fetal interface and that moderate levels of lactonization can regulate redox homeostasis and cell adhesion in the recipient endometrium to promote pregnancy. however, abnormally elevated lactate due to a persistent hypoxic environment may be detrimental to embryonic implantation (14, 15). As the oxygen receptor of cells, hypoxia-inducible factor-1 (HIF-1) plays a major regulatory role in the hypoxic environmental response, and HIF-1 activation in hypoxic environments can lead to reprogramming of energy and nutrient metabolism (16). In contrast, in patients with PE, HIF-1 activation causes CTB invasion dysfunction and incomplete helical artery remodeling in the placenta, leading to excessive lactate accumulation (17, 18). All of the above studies imply that lactate may play a crucial role in the microenvironment of PE patients. Although the work of Yollyseth Medina et al. confirmed that CTB hypoxia in PE patients activates anaerobic glycolysis leading to a functional defect in lactate transport proteins, which in turn leads to excessive lactate accumulation and promotes excessive protein lactation (19–21). However, the effect of hypoxia-induced protein hyperlactivation on PE has never been explored.

Furthermore, mitochondria are the major source of intracellular ROS in hypoxic cells (22). Therefore, mitochondria play an important role in cell fate determination under hypoxia, which is accompanied by increased production of reactive oxygen species (ROS), causing oxidative stress (23), while excess ROS plays a role in cell death key role (24).

A number of studies have examined extensive changes in the human placental transcriptome (25, 26). Liu et al. performed single-cell RNA sequencing from human placenta and identified novel subtypes of trophoblasts, Hofbauer cells, and mesenchymal stromal cells (27, 28). Tsang et al. identified different cell subtypes in human placenta by analyzing more over 24,000 cells, identified different cell subtypes in the human placenta and reconstructed trophoblast differentiation trajectories (29). In addition to this, proteomic assays have rapidly matured, and analysis of proteomics can further yield complementary information combined with transcriptomic data to enhance biomarker discovery (30). However, the changes that occur in various cell types in placental tissues in the presence of hypoxia and the specific molecular mechanisms are not known. Therefore, in this study, we analyzed single-cell sequencing data to comprehensively characterize the molecular and cellular physiopathological abnormal ecology of PE in the hypoxic microenvironment, while combining single-cell transcriptome data with immunofluorescence, and proteomics data to enable the elucidation of PE cellular dynamics mechanisms under complex pathological conditions.

## Materials and Methods

### Data sources

The placenta is a highly heterogeneous organ that is closely associated with adverse pregnancies. Previous bulk sequencing of whole tissues could not reveal the characteristics of individual cells and cell-to-cell interactions. Here, we downloaded the GSE173193 dataset based on the GPL24676 platform from the Gene Expression Omnibus (GEO) database (http://www.ncbi.nlm.nih.gov/geo/), including single-cell sequencing data from placental samples of two PE patients and two normal controls. Sequencing data of placenta samples from the gestational diabetes mellitus group (GDM) and the advanced age group (GL) were excluded from this study.

In addition, this study collected a total of 15 PE patients and 15 placental tissues from a control donor. This study was approved by the Ethics Committee of The Affiliated Yantai Yuhuangding Hospital of Qingdao University and written consent was obtained from all patients or their legal guardians participating in the study.

### Cell lines and culture conditions

HTR-8/SVneo (human immortalized EVT cell line) was purchased and cultured in DMEM/F12 medium (GIBCO, USA), 10% inactivated fetal bovine serum (GIBCO) and 1 % penicillin-streptomycin (GIBCO) at 37 °C and 5 % CO_2_ until the logarithmic growth phase of cells. Cells were inoculated onto six-well culture plates at a rate of 1 × 10^5^ cells/well and incubated in an incubator at 37 °C and 5 % CO_2_. For various experiments under hypoxic conditions, cells were cultured at 37 °C in a humid chamber containing 2 % O_2_ and 5 % CO_2_ (Thermo, 3131, Waltham, Ma, USA).

### Tissue immunofluorescence

PE patients and controls placenta (maternal side) tissue was embedded and sectioned (6 μm thickness), then routinely dewaxed in xylene alcohol, repaired with citric acid and then washed three times with distilled water for 3 min each. the sections were closed with 1% Bovine serum albumin (BSA) for 30 min at room temperature. the primary antibody was diluted with 1% BSA and incubated overnight at 4 degrees. the next day, the sections were washed three times with phosphate buffer solution (PBS) for 3 min each. then the sections were diluted with 1% BSA The fluorescent secondary antibody was incubated for 1 h in a 37°C incubator with closed light, and then the sections were washed three times with PBS for 3 min each time under closed conditions. then the sections were blocked by adding glycerol-diluted hoechst33342 (10 μg/ml) and photographed by laser confocal (Nikon, A1R+). For fluorescent direct-labeled primary antibodies, fluorescently labeled primary antibodies diluted with 1% BSA were added between washing away the secondary antibodies and adding nucleation reagents, and then the sections were washed three times with PBS for 3 min each time. in the case of non-fluorescently labeled secondary antibodies with L-Lactyllysine, 488-labeled goat anti-rabbit secondary antibodies were added after incubating the primary antibodies for another 30 min.

### IGF2BP2 mRNA in situ hybridization and immunofluorescence co-staining

Cy3-tagged IGF2BP2 homo probe sequence: CTTGCCACCT+TTGCCAATCACCCG. RNA in situ hybridization was performed according to the kit instructions, and the main steps were: firstly, paraffin sections were routinely dewaxed, then proteinase K was briefly digested for 5 min in a 37°C incubator, closed and denatured, then the probe was diluted and mixed with biotin according to the kit instructions (i.e. Biotin-probe + SA-Cy3), and then added dropwise to the sections for hybridization, and after the hybridization reaction, the sections were washed with washing solution The sections were washed 15 min × 3 times after the hybridization reaction, and finally the nuclei were stained with 4’,6-diamidino-2-phenylindole (DAPI). For co-staining with other proteins, after probe hybridization and before nucleation, the corresponding fluorescently labeled secondary antibody was added, and since the corresponding protein would be reduced after proteinase K digestion, the antibody concentration was increased and incubated for 30 min at 37°C in the incubator, followed by nucleation.

### Flow cytometry

The tissue was digested with trypsin into single cells, the medium was neutralized and digested and centrifuged to remove the trypsin, the cells were resuspended with PBS (1×10^6^ cells/ml), then fluorophore-labeled flow antibody was added and a blank cell without antibody was set up simultaneously, incubated for 30 min at room temperature in closed incubation on ice, and then the cells were sorted on a flow cytometer (BD, BD Aria II).

PE patients and control CTB, EVT, HTR-8 cells treated with hypoxia and their controls were loaded into 6-well plates at a density of 1×10^4^ cells/ml (three experiments were repeated) and incubated overnight at 37°C at 5% CO_2_ as usual. Add 5 μl of Annexin V (BD, Shanghai) and incubate for 15 min at room temperature, protected from light. Add PI (BD, Shanghai), take a slide to form a small chamber, take the above stained cell suspension from each centrifuge tube and add it to the small chamber, cover the slide and observe the apoptosis by flow cytometry.

### Protein mass spectrometry analysis

The proteins were removed from PE patients after immunoprecipitation from -80 ℃ and lysed by ultrasound. Protein concentration was determined using BCA kit (Beyotime, Shanghai, China). Equal amounts of each sample protein were taken for enzymatic digestion, and the volume was adjusted to the same level with the lysis solution, then DL-Dithiothreitol (DTT, Sigma-Aldrich, Germany) was added to a final concentration of 5 mM, and reduced at 56 ℃ for 30 min. Tetraethylammonium bromide (TEAB, Sigma-Aldrich, Germany) was added to dilute the urea to a concentration below 2 M. Trypsin was added at a ratio of 1:50 (protease:protein, m/m) and digested overnight. The peptides were solubilized with liquid chromatography mobile phase A and separated using an EASY-nLC 1200 UHP system. Mobile phase A was an aqueous solution containing 0.1% formic acid and 2% acetonitrile; mobile phase B was an aqueous solution containing 0.1% formic acid and 90% acetonitrile. The liquid phase gradient settings were: 0-62 min, 4%-23% B; 62-82 min, 23%-35% B; 82-86 min, 35%-80% B; 86-90 min, 80% B, and the flow rate was maintained at 500 nL/min. The peptides were separated by the UHPLC system and then injected into the NSI ion source for ionization and then into the Q Exactive™ HF-X mass spectrometer for analysis. HF-X mass spectrometry was used for analysis. The ion source voltage was set at 2.1 kV and the peptide parent ions and their secondary fragments were detected and analyzed using a high-resolution Orbitrap. The primary mass spectrometry scan range was set to 400 - 1500 m/z, and the scan resolution was set to 120,000; the secondary scan resolution was set to 15,000. The secondary mass spectrometry was performed. In order to improve the effective utilization of the mass spectrum, the automatic gain control (AGC) was set to 5E4, the signal threshold was set to 2.5E5 ions/s, the maximum injection time was set to 40 ms, and the dynamic exclusion time of the tandem mass spectrometry scan was set to 30 s seconds to avoid repeated scanning of the parent ions.

### Cell clustering and differentially expressed genes (DEGs)

The Seurat package (31) was used for cell clustering of scRNA-seq data and to visualize clusters using the UMAP package (32). To define clusters, clusters characterized by similar marker genes were combined into one cell type, and subsequently marker genes for known cell types were used to define the partitioned cell clusters. In addition, the DEGs in each cell type of control donor and PE patient placental tissue were identified by the "FindAllMarkers" function of the Seurat package. Differences associated with adjusted P values, adjusted P<0.05, were considered significant.

### Gene Regulatory Network (GRN)

The Single Cell Regulatory Network Inference and Clustering (SCENIC) algorithm has been developed to evaluate regulatory network analysis associated with transcription factors (TFs) and to discover regulators (i.e., TFs and their target genes) in individual cells. To reconstruct gene regulatory networks from scRNAseq data in control donors and PE patients, we performed SCENIC analysis, which uses co-expression modules between TFs and candidate target genes, as well as a database of DNA binding patterns of TFs to infer important gene regulation of transcription factors (33, 34). Regulon modules based on regulon crosstalk (correlation between regulons and regulators) were determined by the connection specificity index (CSI), which ranks the importance of regulons and mitigates the effect of non-specific interactions, and visualized based on the R package ComplexHeatmap (35).

### Pseudo-Time Analysis

In this study, single-cell trajectory analysis of cell subtypes was performed as needed. The R package Monocle 3 was applied to sort cells along the trajectory in pseudo-time (https://cole-trapnell-lab.github.io/monocle3) (36). After clustering the cells using the above method, the dimensionality was reduced and the results were visualized using the UMAP method. Subsequently, cells were sorted according to their progression through the developmental program. Monocle measured this progression in pseudo-time.

### Functional enrichment analysis

Gene ontology (GO) enrichment of cell clusters and Kyoto Encyclopedia of Genes and Genomes (KEGG) pathway analysis were used to determine the biological significance of each cell type. GO and KEGG pathway analysis was performed using the clusterProfiler package for marker genes for each cell subpopulation (37). In addition, Gene Set Enrichment Analysis (GSEA) was performed on DEGs (38) using c2.cp.kegg.v7.0.symbols.gmt from the MsigDB database (39) as a background set. GSEA was performed using the clusterProfiler package to perform and P<0.05 was considered significant.

### Analysis of intercellular communication

To further explore the interactions between different cell subpopulations, cellular communication analysis was performed using the R language iTALK package. iTALK identifies genes that are highly or differentially expressed in cell clusters and matches and pairs these genes through a receptor-ligand database to find important cellular intercellular communication events. Receptor-ligand interactions were determined using the STRING database of protein-protein interactions (40).

### Molecular docking

The three-dimensional structural (41) of the lactate molecule was downloaded from PubChem (https://pubchem.ncbi.nlm.nih.gov/), a public repository for biological activity data of small molecules and RNAi reagents. And the three-dimensional structure of PTBP1 protein was downloaded from the Protein Data Bank (https://www.rcsb.org) (42), and the binding sites and energy predictions of the lactate molecule and PTBP1 protein were evaluated by Autodock (43). In addition, Hex 8.0.0 (44) was used to predict the binding sites and binding potential of PTBP1 protein to IGF2BP2 RNA molecules before and after lactylation. Pymol (45) was applied to visualize the docking model. Docking energy less than 0 indicates that the two have binding potential, and the lower the energy the higher the binding potential.

### Data Analysis and Statistics

In this study, all analyses were performed based on the Bioinforcloud platform (http://www.bioinforcloud.org.cn).

## Results

### Global single-cell landscape of the PE patients placenta

The analytical flow of this study is shown in **Figure 1A**. Based on single-cell sequencing, we constructed a global single-cell landscape of PE patients placenta involving 46 cell clusters of 26,416 cells. These cell clusters were identified into 12 cell types based on known markers, including Hofbauer, cytotrophoblasts (CTB), neutrophils, extravillous trophoblasts (EVT), blood cells (BC), maternal uterine dendritic cells (DC), syncytiotrophoblasts (STB), eosinophils, B cells, T cells, placental endothelial cells (En), and ecdysteroid cells (Dec) (**Figure 1B**). Each cell type showed specific expression of cell type markers (**Figure 1C**). Specifically, multiplex immunofluorescence experiments revealed that EVTs from PE patients placenta had less vascular endothelial growth factor receptor 1 (FLT1) relative to controls placenta (**Figure 1D**), whereas EVTs from the PE group had higher levels of lactic acidification modifications (**Figure 1E**). Overall, we constructed a global single-cell atlas of PE and control placenta with precise cell type annotation, depicted cellular ecological differences between PEs and controls placenta, and experimentally validated the levels of abnormal angiogenesis and lactic acidification modifications in EVTs.

**Figure 1.**
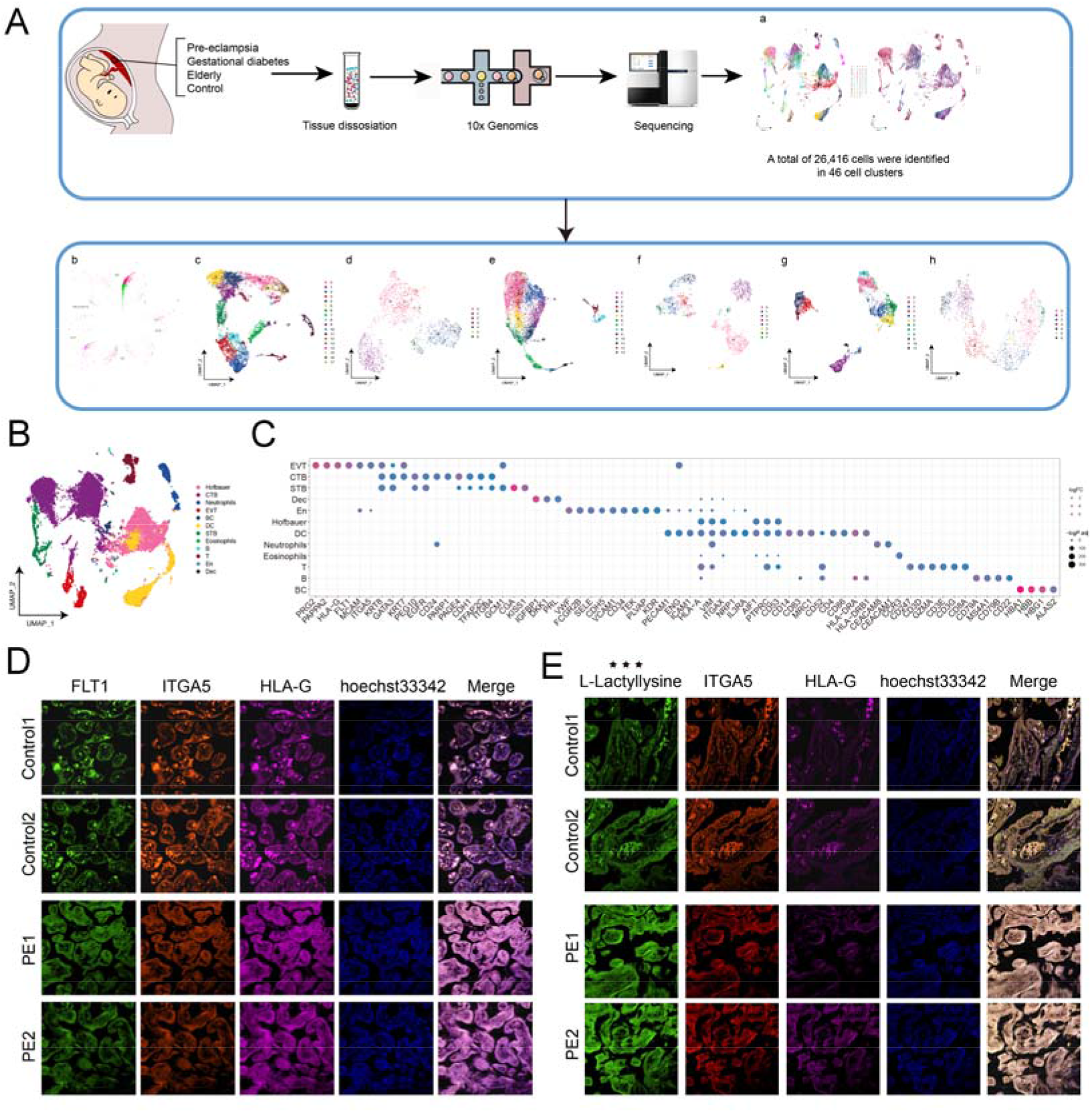
Global single-cell landscape of PE patients placenta. A. Study flow of this work. A global single-cell landscape of PE patients placenta was constructed based on single-cell technology, capturing a total of 26,416 cells from 46 cell clusters. B. Single-cell atlas mapping cell types. including fetal macrophages (Hofbauer), cytotrophoblasts (CTB), Neutrophils, extravillous trophoblasts (EVT), blood cells (BC), maternal uterine dendritic cells (DC), syncytiotrophoblasts (STB), Eosinophlis, B cells, T cells, placental endothelial cells (En), and ecdysteroid cells (Dec). C. Bubble plots showing cellular markers guiding cell annotation. D. Multiplex immunofluorescence plots showing PEs and controls extravillous trophoblast (EVT) and endothelial landscapes. Highlights are erythrocytes, compared to more vascular/hematocrit in the controls and less in the PEs. E. Multiplex immunofluorescence map demonstrating the level of lactonization modification of EVT in the PEs and controls.

### CTB cells in PE patients significantly inhibit cell cycle activation of apoptotic signaling

During human pregnancy, placenta-derived CTB can differentiate into EVT subpopulations involved in placental angiogenesis, which in turn play a key role in normal fetal growth and development. Therefore, dysfunction of these subpopulations may be an initiating risk factor mediating the development of PE. To this end, CTB cells were extracted for subpopulation identification to explore the possible mechanisms involved in these subpopulations in PE. A total of 14 subpopulations were obtained through the identification of CTB subpopulations (**Figure 2A**). Further comparison of cell abundance revealed that the proportion of cellular CTB_CA8 subpopulation was significantly increased and the proportion of CTB_SGMS2 subpopulation was significantly decreased in PE, and these two subpopulations were identified as specific CTB subpopulations (**Figure 2B**), and the expression of CA8 and SGMS2 was subsequently mapped in the single cell profiles of CTB subpopulations (**Figure 2C**). To further explore the mechanism of CTB subpopulation involvement in PE, we extracted marker genes of CTB_CA8 subpopulation and CTB_SGMS2 subpopulation for enrichment analysis. The results showed that the HIF-1 signaling pathway, apoptosis and TNF signaling pathway were significantly enriched in the CTB_CA8 subpopulation, while the cell cycle was significantly enriched in the CTB_SGMS2 subpopulation (**Figure 2D**). The KEGG pathway map further revealed that the cell cycle activity of CTB_CA8 cell subpopulation was reduced, while HIF-1 signaling pathway, apoptosis and necroptosis pathway were significantly activated (**Figure 2E**). Apoptosis and necroptosis were found to be more pronounced in PE by flow cytometry experiments (**Figure 2F**). Subsequent proposed chronological analysis revealed the developmental trajectory of CTB subpopulations, and both specific CTB subpopulations were in an early developmental position (**Figure 2G**). GRN revealed that CTB subpopulation marker genes were divided into 4 modules (**Figure 2H**), which were mainly regulated by TFs such as HEY2, HOXB6 and ELF4 (**Figure 2I**). In conclusion, we identified significant cell cycle suppression and activation of apoptotic signaling by CTB cells in PE, and inferred the origin and trajectory of CTB cell pathogenesis and evolution, and constructed a GRN of CTB cells in the development and progression of PE.

**Figure 2.**
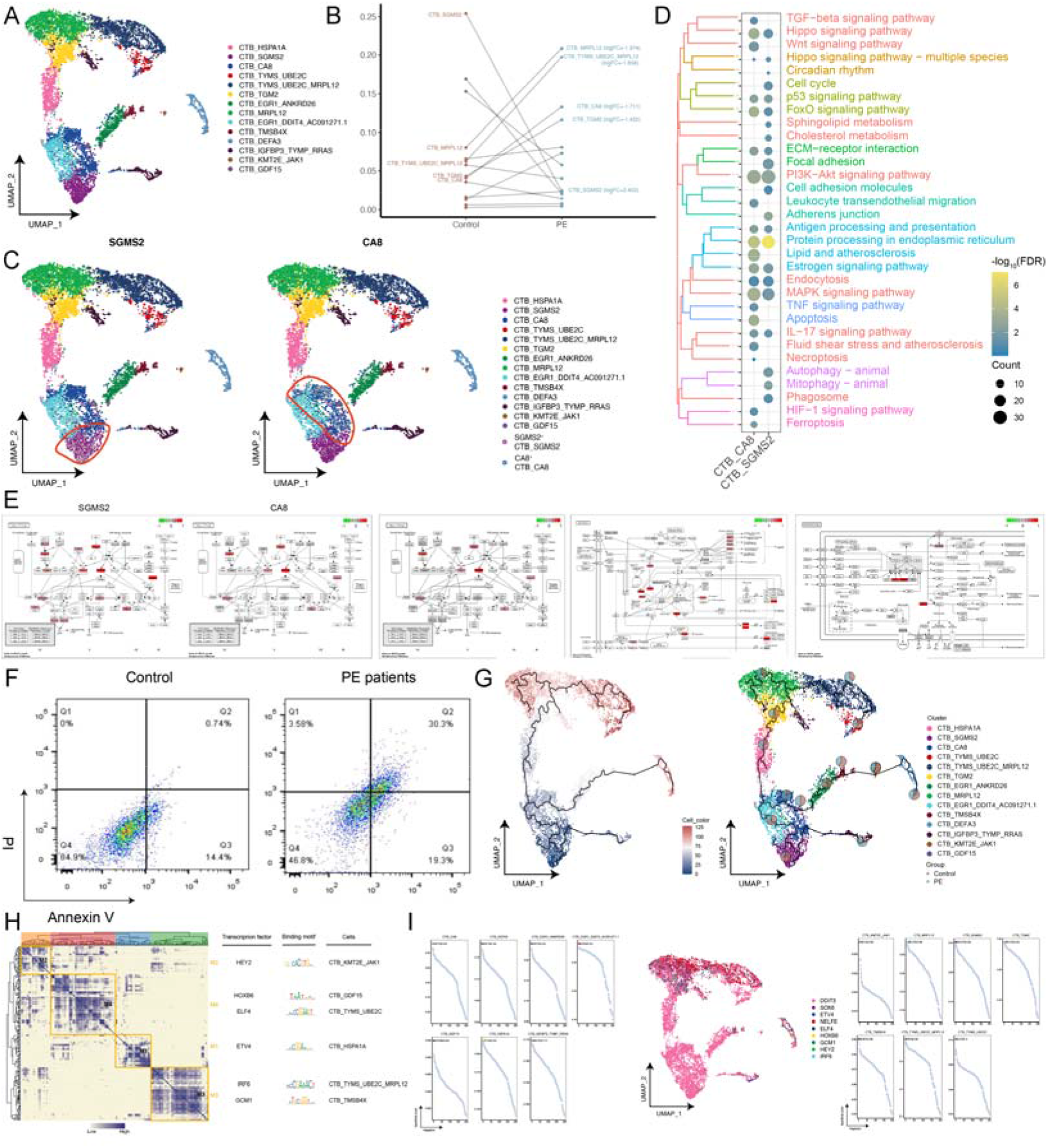
Identification of CTB subpopulations in PE patients and exploration of the mechanisms involved. A. Single-cell atlas demonstrating CTB subpopulations. B. Changes in the ecological composition of CTB in Controls-PEs. C. Markers of PE patients-specific CTB. D. Biological pathways significantly involved in CTB_CA8 and CTB_SGMS2 cell subpopulations. E. Activation or inhibition of cell cycle, HIF-1 signaling pathway, apoptosis and necroptosis pathways in CTB_CA8 cell subpopulations. F. Flow cytometry experiments to verify apoptosis and necroptosis in CTB from PE patients and controls. G. Pseudotime analysis of CTB subpopulations in PE patients. H. GRN of CTB cell subpopulations in PE patients. I. CTB subpopulation-specific transcription factors in PE patients.

### PE patients-specific EVT_CSNK2B cell subpopulation and control EVT_FUCA1_FLNB cell subpopulation

Pregnancy is a complex and delicate process, and after blastocyst implantation in early gestation, placental EVT cells further invade, migrate and differentiate towards the metaplastic and myometrial vasculature of the uterus, initiating and completing vascular remodeling, which is a critical part of establishing maternal-fetal circulation. Therefore, we also extracted EVT cells for subpopulation identification and captured a total of 6 EVT subpopulations (**Figure 3A**). By comparing the cell abundance, it was found that the proportion of EVT_CSNK2B subpopulation was significantly increased in PE compared with the control, while the proportion of EVT_FUCA1_FLNB subpopulation was significantly decreased (**Figure 3B**). Enrichment analysis revealed that markers of EVT_CSNK2B subpopulation were significantly involved in pathways such as necroptosis, iron death, and HIF-1 signaling pathway, while markers of EVT_FUCA1_FLNB subpopulation were significantly involved in pathways such as adhesive junctions, adherent spots, and tight junctions (**Figure 3C**). Subsequently, the expression of CSNK2B and FUCA1 was mapped in the single-cell profiles of EVT subpopulations, while immunofluorescence showed low FLNB expression and high CSNK2B expression in EVT cells with PE (**Figure 3D**), which was consistent with the results of the raw signal analysis (**Figure 3B**). The GRN of EVT cell subpopulations suggested that the markers of EVT cells were divided into 5 modules in total (**Figure 3F**) and regulated by MAFG, SRF, KLF7, and ETV4 TFs (**Figure 3G**). In conclusion, we identified subpopulations of EVT cells in PE and the pathways their participate in, inferred the origin and trajectory of the evolution of EVT cell pathology, and constructed a GRN of EVT cells in the development and progression of PE.

**Figure 3.**
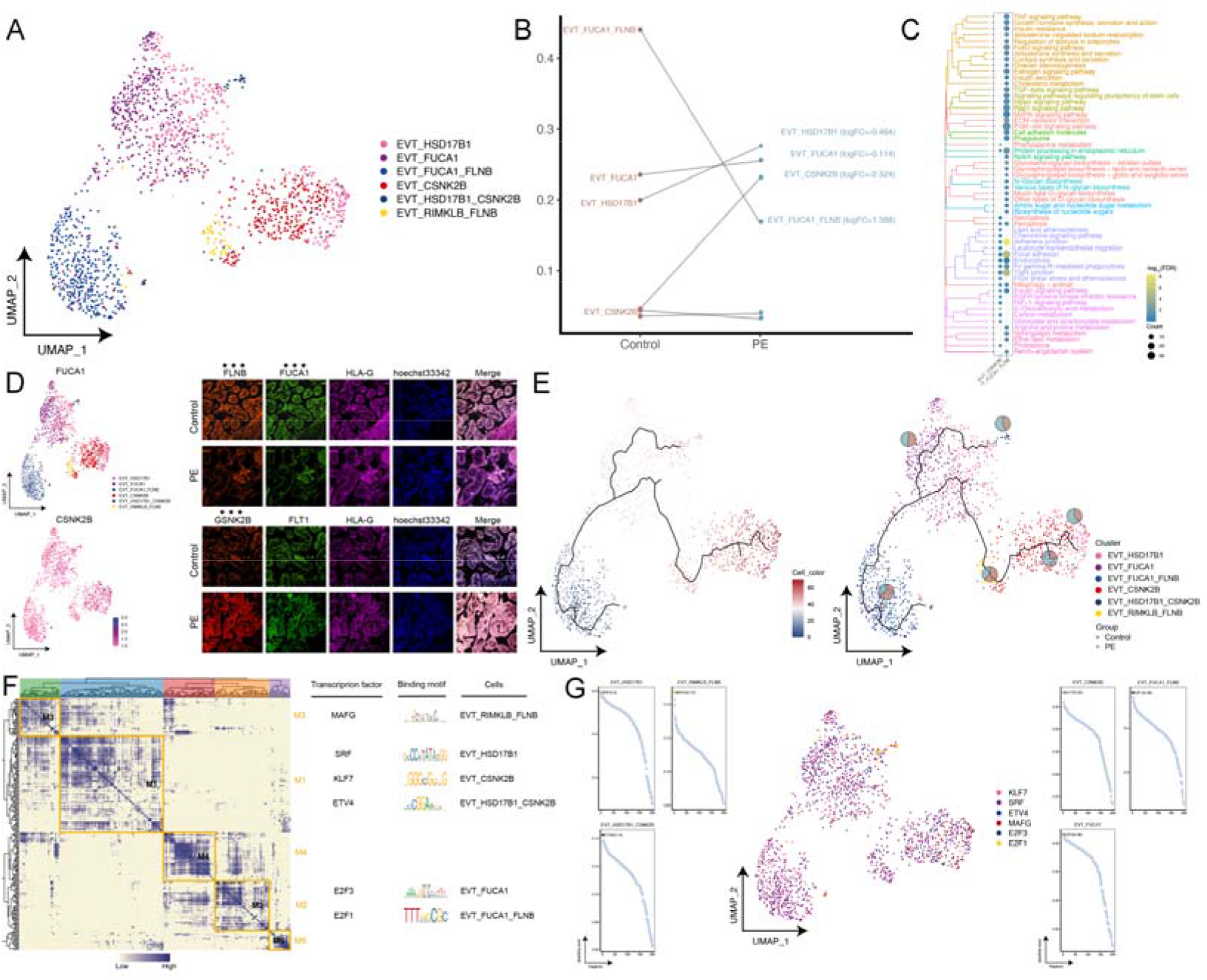
Identification of EVT subpopulations in PE and exploration of the mechanisms involved. A. Single-cell atlas demonstrating EVT cell subpopulations. B. Dotted line graph demonstrating EVT cell ecology of control-PE. C. Biological pathways significantly involved in EVT_CSNK2B and EVT_FUCA1_FLNB cell subsets. D. Series of single-cell atlas-multiple immunofluorescence plots demonstrating labeling of PE patients-specific EVT cell subpopulations. E. Pseudotime analysis of EVT cell clusters in PE patients. F. GRN of EVT cell subpopulations of PE patients. G. EVT cell subpopulation-specific transcription factors in PE patients.

### Inhibition of IGF2BP2 expression causes defective mitochondrial autophagy and adherent patch signaling in EVT of PE patients

As a specific subpopulation of EVT, the EVT_CSNK2B subpopulation may play an important role in the development of PE. Therefore, to further explore the potential mechanisms involved in the development of PE, we performed GSEA on the EVT_CSNK2B subpopulation. The results showed that mitochondrial autophagy, adherent spots and associated metabolic signals were inhibited in the EVT_CSNK2B subpopulation (**Figure 4A**). We then constructed a comprehensive regulatory network of upstream regulators (IGF2BP2, PLXNB2, TCF7L2, MBNL2, MYC, RBFOX2, MLLT1, NEAT1) on mitochondrial autophagy, adherent spots and associated metabolic signaling in the EVT_CSNK2B cell subpopulation (**Figure 4B**). Intriguingly, IGF2BP2 is an N6-methyladenosine (m^6^A)-associated gene that enhances mRNA stability and promotes translation as an m^6^A reader protein. Therefore, we mapped its expression in single-cell profiles of EVT subpopulations and found that IGF2BP2 was widely expressed in the dominant subpopulation of control pregnant women, EVT_FUCA1_FLNB, yet significantly reduced in the PE-specific EVT_CSNK2B cell subpopulation (**Figure 4C**). Furthermore, multiplex immunofluorescence experiments confirmed that IGF2BP2 was lowly expressed in EVT cells from placenta in the PE patients compared to controls (**Figure 4D, E**). Briefly, inhibition of IGF2BP2 expression in PE causes defective mitochondrial autophagy and adherent spot signaling in EVT.

**Figure 4.**
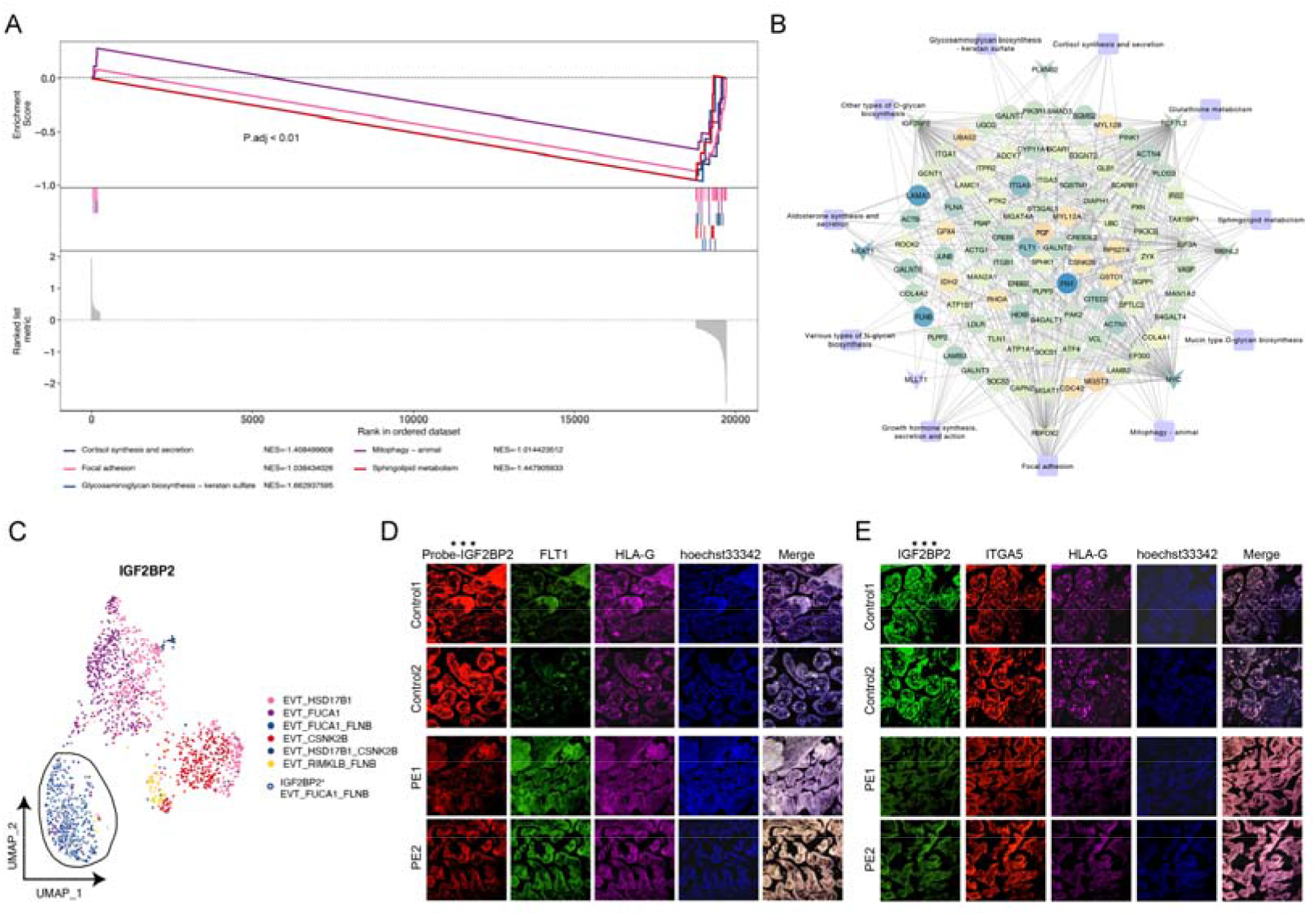
Inhibition of IGF2BP2 expression causes defective mitochondrial autophagy and adherent patch signaling in EVT of PE patients. A. EVT_CSNK2B cell subpopulation significantly inhibits mitochondrial autophagy, adherent spots and associated metabolic signaling. B. Network diagram demonstrating the regulation of mitochondrial autophagy, adherent spots and related metabolic signaling by upstream regulators in EVT_CSNK2B cell subpopulation. C. Single cell profiles demonstrating the expression of IGF2BP2. D-E. Multiplex immunofluorescence assays to verify the RNA and protein abundance of IGF2BP2 in EVT cells from PE patients and controls.

### Hypoxia promotes PTBP1 lactonization leading to PTBP1 inactivation inhibiting RNA processing and synthesis of IGF2BP2 mediating the disease process of PE

However, it is puzzling what causes the suppression of RNA expression of IGF2BP2 in PE. To this end, we identified the core molecule regulating IGF2BP2 expression PTBP1. Notably, there was no significant difference in the protein abundance of PTBP1 in EVT cells from PE patients in multiplex immunofluorescence assays (**Figure 5A**), and there was no binding of PTBP1 protein and IGF2BP2 probe in EVT cells from PE patients with hyperlactation, but there was binding in controls, suggesting that PTBP1 protein in placental EVT from PE patients had a weaker ability to bind RNA molecules of IGF2BP2 (**Figure 5B**). Furthermore, the binding potential of PTBP1 and lactic acid small molecules was identified by molecular docking (**Figure 5C**), indicating that PTBP1 protein is capable of lactonization and that the structure of lactonized PTBP1 protein is disrupted. In addition, the post-lactated PTBP1 protein had a much lower RNA binding potential to IGF2BP2 (**Figure 5D**).

**Figure 5.**
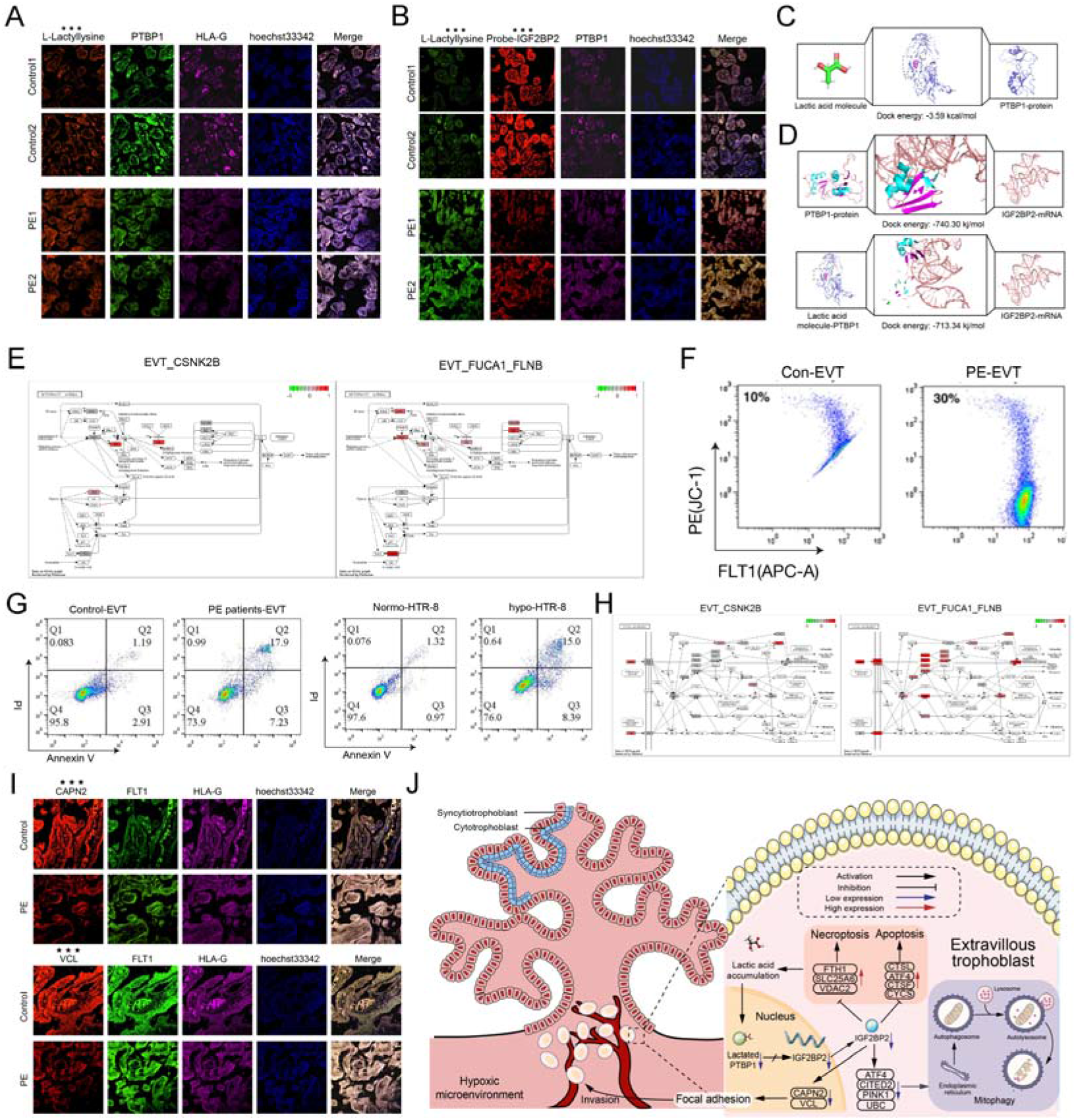
Hypoxia promotes PTBP1 lactylation leading to PTBP1 inactivation inhibiting the processing of RNA synthesis of IGF2BP2 mediating the disease process of PE patients. A. Multiplex immunofluorescence assay to verify the protein abundance of PTBP1 in EVT from PE patients and controls. B. Multiplex immunofluorescence assay to verify the ability of PTBP1 protein to bind RNA molecules of IGF2BP2 in PE patients (hyperlactylated) and control (hypolactylated) EVT. C. Molecular docking predicts the binding site and potential of PTBP1 lactylation modification. D. Molecular docking predicts the binding sites and potential of PTBP1 protein to IGF2BP2 RNA molecules before and after lactylation. E. Activation or inhibition of mitochondrial autophagy in EVT_CSNK2B and EVT_FUCA1_FLNB cell subpopulations. F. Flow cytometry validation of mitochondrial autophagy in EVT from PE patients and controls. G. Flow cytometry validation of apoptosis and necrotizing apoptosis in PE patients, control EVT and HTR-8 cell lines before and after hypoxia treatment. H. Activation or inhibition of adherent spot signaling in EVT_CSNK2B and EVT_FUCA1_FLNB cell subpopulations. I. Multiplex immunofluorescence assay to verify the activation or inhibition of adherent spot signaling in EVT from PE patients and controls. J. Under normal conditions, PTBP1 protein promotes the processing and synthesis of IGF2BP2, which in turn promotes the mitochondrial autophagy and adherent patch pathway and inhibits the apoptotic and necroptotic pathways in EVT. However, lactylation of PTBP1 protein in EVT under hypoxic microenvironment leads to its inactivation, which in turn blocks the processing and synthesis of IGF2BP2 RNA, and the suppressed expression of IGF2BP2 drives the disease process of PE through two pathways. Pathway 1: Promotes damage depletion of mitochondria and inhibits mitochondrial autophagy, causing accumulation of damaged mitochondria, inducing apoptosis and necroptosis of EVT while blocking lactate metabolism and exacerbating lactate accumulation, forming a feedback vicious inner circle; Pathway 2: Inhibits the adhesive patch signaling pathway of EVT, weakening the invasive ability of EVT and leading to insufficient reconstruction of placental spiral vasodilation, forming a feedback vicious external circulation.

The effect of suppression of RNA expression of IGF2BP2 on PE has not been clarified. Therefore, we explored in depth the mechanisms involved in IGF2BP2 inhibition. First, we found that the mitochondrial autophagic pathway in EVT_CSNK2B cell subpopulation was significantly activated (**Figure 5E**). Flow cytometry showed that mitochondrial autophagy was inhibited after placental EVT cells were treated with hypoxia (**Figure 5**F), and not only that, the cells showed early apoptosis and necrotizing apoptosis in EVT cells of PE and hypoxic HTR-8 cell line (**Figure 5**G).

Since significant activation of adherent spot signals is a major manifestation of cell migration and invasion, we also explored the activation of invasion-related signals such as adherent spots in EVT subpopulations after IGF2BP2 inhibition. We found that adherent patch signaling was significantly suppressed in the EVT_CSNK2B cell subpopulation (**Figure 5H**), and in addition, multiple immunofluorescence experiments confirmed that the expression of the key molecule of adherent patch signaling (CAPN2) was suppressed in EVT cells from PE patients (**Figure 5I**).

In summary, our results show that lactylation of PTBP1 in EVT cells under hypoxic microenvironment leads to its inactivation, inhibits RNA processing synthesis of IGF2BP2, promotes damage depletion of mitochondria and inhibits mitochondrial autophagy, causing accumulation of damaged mitochondria, inducing apoptosis and necrotic apoptosis of EVT cells while blocking lactate metabolism and intensifying lactate accumulation, forming a feedback It also inhibits the EVT adhesion spot signaling pathway and weakens the invasive ability of EVT, leading to insufficient reconstruction of placental spiral vasodilation and forming a feedback vicious external loop. The continued activation of the malignant internal and external dual circulation system in the hypoxic microenvironment drives the disease process of PE (**Figure 5J**).

### Microenvironmental immune cell ecology of the placenta in PE

During different periods of pregnancy, the immune microenvironment at the maternal-fetal interface exhibits a dynamic balance, which facilitates the establishment and maintenance of pregnancy as well as fetal growth and development. To this end, we extracted immune cells from PE patients placenta for subpopulation identification to explore the microenvironmental immune cell ecology of PE patients placenta. A total of 11 subpopulations were captured by subpopulation identification of the indicated Hofbauer cells (**Figure 6A**), and the distinctive markers of these subpopulations are shown (**Figure 6C**). Among them, the Hofbauer_ALPL_NBN subpopulation was significantly reduced in proportion in PE, while the Hofbauer_BTG3 subpopulation was significantly increased (**Figure 6B**). For Neutrophils, a total of 5 subgroups were captured, of which the Neutrophils_S100A12 subgroup had a significantly higher proportion in PE, while the markers of these subgroups are shown (**Figure 6D-F**). For DCs, a total of 5 subpopulations were captured (**Figure 6G**), and the proportion of DC_NEU1_IGF1_CBLB_PDE2A_BCL2L1 subpopulation was significantly elevated in PE (**Figure 6H-I**). For T cells, a total of 7 subpopulations were captured (**Figure 6J**), with the proportion of T_CHPT1 subpopulation and T_LRRN3 subpopulation significantly elevated in PE (**Figure 6K-L**). Overall, by exploring the differences in the composition of these immune cell subpopulations in controls and PE patients placenta, we preliminarily characterized the immune cell ecology of the microenvironment in PE.

**Figure 6.**
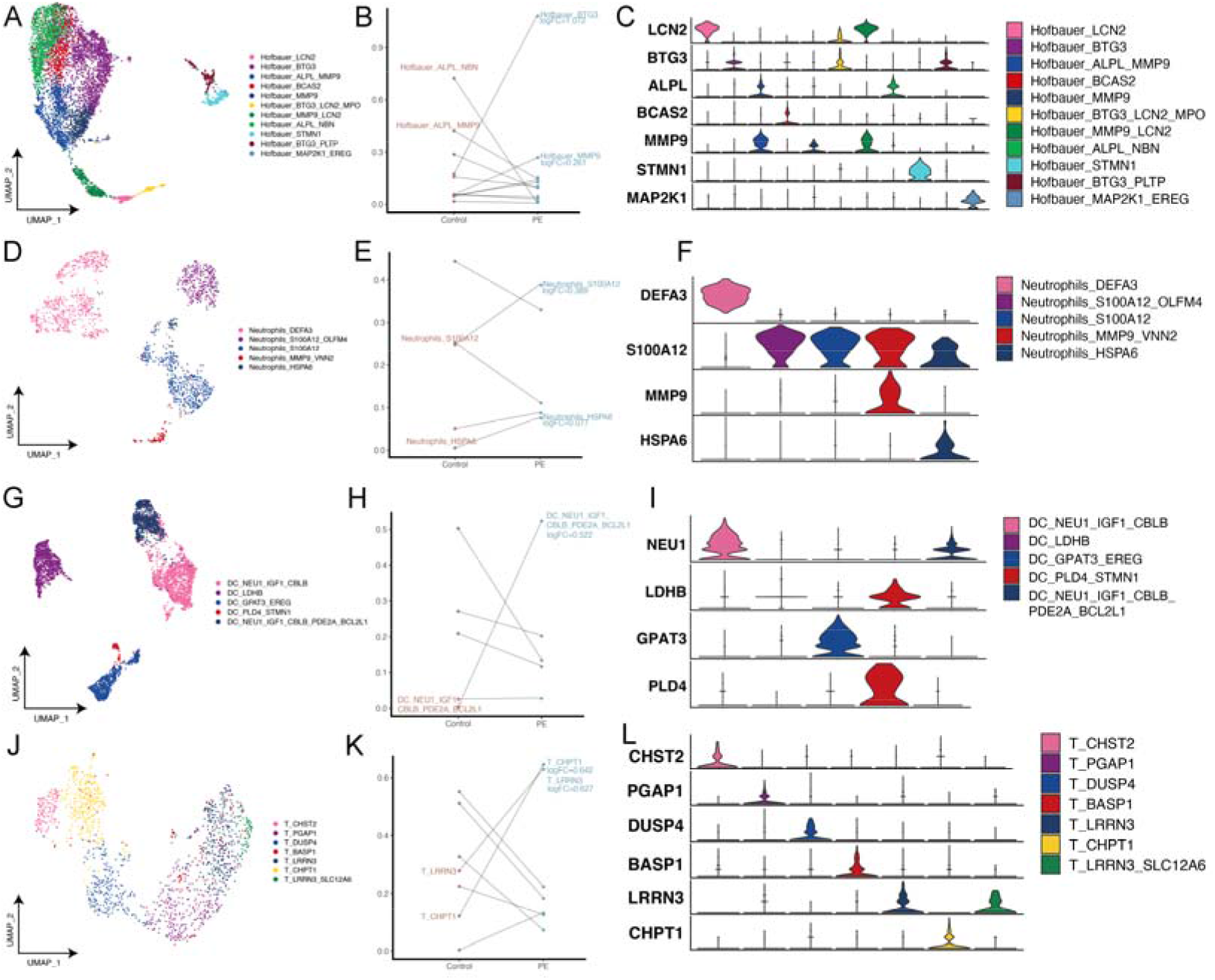
Exploration of the microenvironmental immune cell ecology of the placenta of PE patients. A. Single-cell atlas demonstrating Hofbauer cell subpopulations. B. Variation in the ecological composition of Hofbauer cells in PEs-Controls. C. Violin diagram demonstrating markers of PE-specific Hofbauer cell subpopulations. D. Single-cell plots demonstrating Neutrophils cell subpopulations. E. Changes in the ecological composition of Neutrophils cells in PEs-Controls. F. Violin diagram showing markers of PE-specific Neutrophils cell subpopulations. G. Single-cell plots demonstrating DC cell subpopulations. H. Dotted line diagram demonstrating changes in the ecological composition of DC cells in PEs-Controls. I. Violin diagram showing markers of PE-specific DC cell subpopulations. J. Single-cell mapping demonstrating T cell subpopulations. K. Variation in the ecological composition of T cells in PEs-Controls. L. Violin diagram demonstrating markers of PE-specific T cell subpopulations.

### Abnormal metabolic signaling of PE patients-specific EVT subpopulations interacts with microenvironmental immune cells to activate metabolic inflammation

Previous studies have shown that each week of pregnancy involves a plethora of physiological changes and metabolic adaptations, and that even small deviations from normal norms may have adverse consequences at different stages of pregnancy.

Therefore, we explored the interaction of abnormal metabolic signaling in EVT subpopulations with microenvironmental immune cells. Based on KEGG pathway maps, we found that the EVT_CSNK2B cell subpopulation significantly promoted glutathione (anti-inflammatory metabolite) metabolism, inhibited sphingolipid (pro-inflammatory metabolite) metabolism, and suppressed the biosynthesis of various glycans (anti-inflammatory metabolites) (**Figure 7A**). The prevalence of interactions between the EVT_CSNK2B subpopulation of PE and immune cell subpopulations was revealed by cell communication analysis (**Figure 7B**), and the genes for these interactions were further demonstrated (**Figure 7C**). GSEA showed that immune cell clusters of PE, including Hofbauer cells, Neutrophils cells, DCs, and T cells, generally significantly activated immune inflammation-related pathways (**Figure 7D-G**). Taken together, these results suggest an interaction between abnormal metabolic signaling of PE patients-specific EVT subpopulations and microenvironmental immune cells, which activates metabolic inflammation.

**Figure 7.**
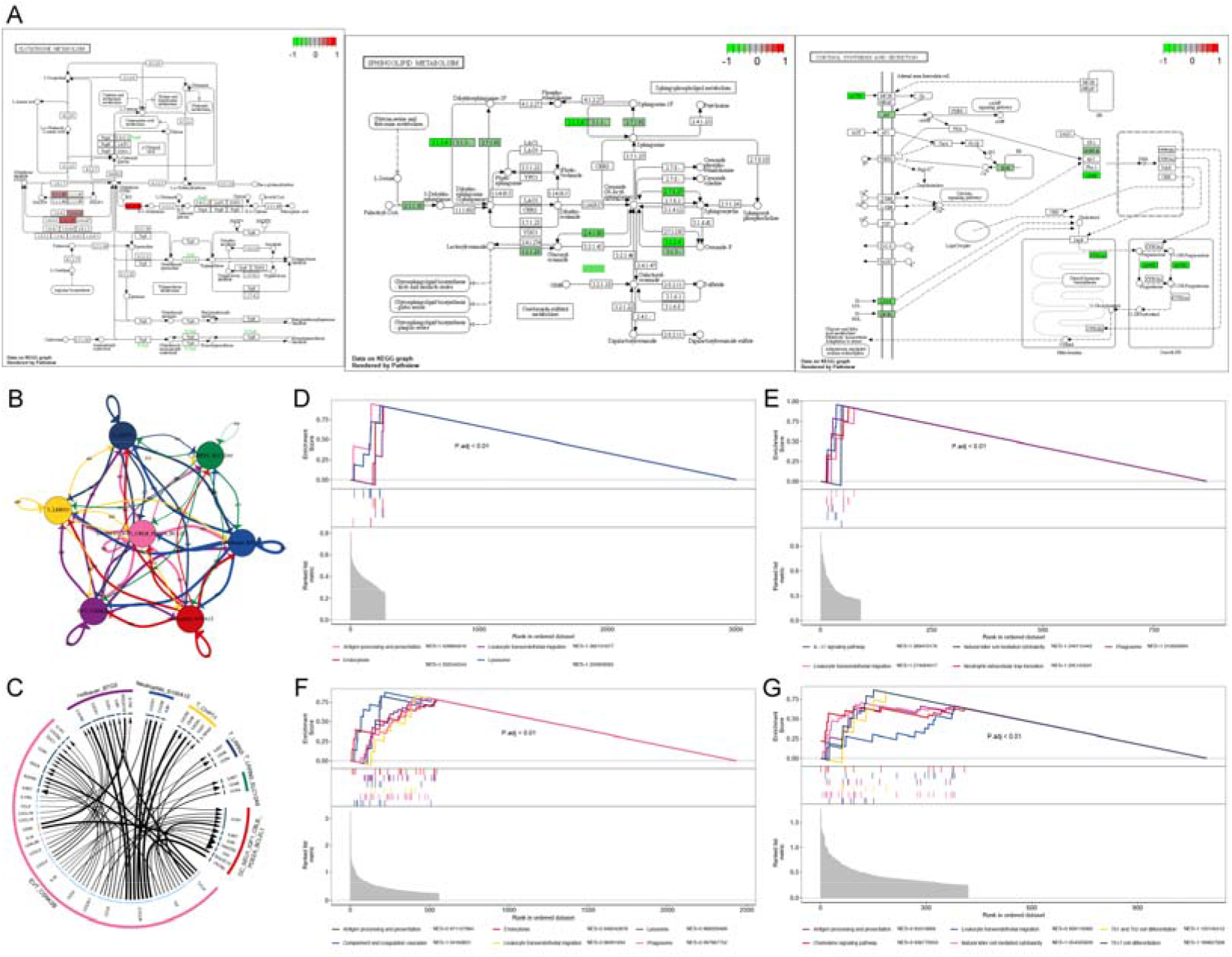
Interaction of abnormal metabolic signaling of PE patients-specific EVT subpopulations with microenvironmental immune cells is explored. A. EVT_CSNK2B cell subpopulation significantly promotes glutathione (anti-inflammatory metabolite) metabolism, inhibits sphingolipid (pro-inflammatory metabolite) metabolism, and suppresses biosynthesis of various glycans (anti-inflammatory metabolites). B and C. Overview of communication events between PE patients-specific EVT_CSNK2B subpopulations and immune cell subpopulations. D. Immunoinflammatory pathways significantly activated by PE patient-specific Hofbauer cells. E. Immunoinflammatory pathways significantly activated by PE patients-specific Neutrophils cells. F. Immunoinflammatory pathway significantly activated by PE patients-specific DC cells. G. Immunoinflammatory pathways significantly activated by PE patients-specific T cells.

## Discussion

This study provides a comprehensive and in-depth dissection of cellular dynamics in the placenta of PE patients from single-cell sequencing data, thus enabling the elucidation of the abnormal molecular and cytopathological ecology of protein hyperlactation-mediated PE in the hypoxic microenvironment under complex pathological conditions.

In the global single-cell ecosystem of PE, we found an increased proportion of carbonic anhydrase 8 (CA8)-positive CTBs (CTB_CA8) in almost all patients, accompanied by reduced cell cycle activity and significant activation of the HIF-1 pathway and apoptotic cell death pathways.CTB cells in the placenta of PE patients undergo excessive cell death, including apoptosis, and if left unchecked, these processes lead to apoptosis sweeping throughout the syncytium eventually stimulating extensive death at this fundamental interface of maternal-fetal exchange (46, 47). Thus, maintaining homeostasis of the trophoblastic hypoxic microenvironment is essential to slow or inhibit PE progression. In addition, studies have captured an abnormally increased subpopulation of CSNK2B-positive EVT in the placenta of PE patients. Based on comprehensive regulatory network analysis of this subpopulation, m^6^A reading protein insulin-like growth factor 2 mRNA binding protein 2 (IGF2BP2) was identified as a key regulator involved in mitochondrial autophagy and adherent plaque signaling pathways. IGF2BP2 is one member of the IGF2BP family that has previously been regarded as oncofetal, as its members were originally discovered in developing embryos (48). IGF2BP2 is an important gene associated with type 2 diabetes (49). Another IGF2BP family member, IGF2BP3, was found to stimulate trophoblast cell invasion and migration. In addition, in recent studies, IGF2BP2 was confirmed which trophoblast cells invade and migrate. However, no study has yet investigated the invasion and migration of IGF2BP2 at the single cell level in PE (50, 51). It is well known that post-transcriptional modifications of messenger RNAs (mRNAs), among which m^6^A is the most abundant internal RNA modification, regulate genes expression by influencing mRNA splicing, stability, translocation and translation (52). Moreover, m6A modification exerts its biological functions by “readers”: YTH (YT521-B homology) domain proteins including YTHDC1-2 and the YTH-family proteins YTHDF1–3 as well as insulin-like growth factor 2 mRNA binding proteins IGF2BP1-3 (53). Among these m^6^A readers, IGF2BP2 binds RNA via its six characteristic RNA-binding domains, containing two RNA recognition motifs (RRM1 and RRM2) and four K Homology (KH) domains (KH1-KH4) (54).

In addition, we identified a core regulator, polypyrimidine bundle-binding protein 1 (PTBP1), in a study of lactonization in a CSNK2B-positive EVT subpopulation. Previous studies have demonstrated that PTBP1 is a known regulator of post-transcriptional gene expression with representative roles in controlling mRNA splicing, translation, stability and localization, and more strikingly, recent studies have identified its involvement in glycolytic processes. PTBP1 is a key determinant of pyruvate kinase isoform 2 in PKM embryos and is used to promote aerobic glycolysis (55–58). PTBP1 is closely related to the initial formation of the embryo, and PTBP1 serves as a factor essential for early embryonic development, especially because it is necessary for rapid cell division and is essential for embryonic stem cell proliferation (59). However, its potential role and specific molecular mechanisms have not been reported in PE. Our study concluded that the mechanism of IGF2BP2 low expression contributes to the poor internal and external circulation of PE. First, PTBP1 lactonization mediates its inactivation and drives IGF2BP2 low expression, which in turn inhibits mitochondrial autophagy and leads to the continuous accumulation of damaged mitochondria, blocking cellular lactate metabolism for energy while promoting EVT apoptosis and further accumulation of lactate, resulting in a feedback poor internal circulation. Second, PTBP1 lactation-mediated IGF2BP2 low expression also significantly inhibits the adherent plaque signaling pathway, which significantly reduces the invasive capacity of EVT and becomes a persistent obstacle to spiral artery reconstruction, further exacerbating the placental ischemic and hypoxic microenvironment and creating a feedback adverse external circulation. A recent study by Vangrieken P found signs of increased oxidative stress and mitochondrial dysfunction in the placenta of PE, suggesting that mitochondria could be a potential novel therapeutic target in PE (60). In previous studies, mitochondrial autophagy was considered as a cellular defense that removes excess mitochondrial fragments in PE (61). However, in the present study, mitochondrial autophagy was found to be inhibited in EVT cells of PE patients. The HIF-1 signaling pathway was significantly activated, suggesting that hypoxia induces oxidative stress in PE. Furthermore, migration/invasion of EVT cells is tightly regulated by some adhesion molecules, mainly in an autocrine or paracrine manner at the fetal-maternal interface in human pregnancy (62). In the present study, the expression of genes related to adhesive patch signaling was found to be significantly downregulated in PE, suggesting that m^6^A methylated IGF2BP2 reading defects inhibit adhesive patch signaling and reduce the invasive capacity of EVT cells.

Analysis of PE patients placental microenvironmental immune cells in combination with EVT showed that the microenvironmental immune cell function in PE is mainly focused on immune response, suggesting that placental immune function is altered in PE patients. On the one hand, EVT requires remodeling of the uterine spiral arteries, as well as invasion of the metaplastic tissue of the uterus and anchoring of the placenta to the uterine wall. During these processes, different types of microenvironmental immune cells are encountered (63). In order to adapt to each other, a rational immune regulation is necessary.

In summary, our study establishes a large-scale single-cell transcriptome ecological landscape of normal and PE patients placenta and demonstrates changes in cellular dynamics in PE patients placenta, elucidating the abnormal ecological balance of molecular and cellular physiopathology mediated by protein hyperlactation in PE in the hypoxic microenvironment. This opens an avenue to resolve cellular dynamics and aberrations in the complex biological system of PE and to establish a molecular diagnosis of PE. However, the current study still has some limitations. Firstly, our results should be validated in a large-scale study due to the insufficient sample size. Second, the interactions between these abnormal cells in PE deserve further investigation.

## Data Availability

The datasets analyzed during the current study are available in the GEO database

https://www.ncbi.nlm.nih.gov/geo/

## Data Availability Statement

The datasets analyzed during the current study are available in the GEO database (https://www.ncbi.nlm.nih.gov/geo/).

## Acknowledgments

Not applicable.

## Author Contributions

Hongmei Qu, Xiaoyan Li and Qian Li analyzed the data and drafted the manuscript. Linsong Mu, Yanfen Zou and Yongli Chu designed the study and revised the manuscript. Yan Feng, Xiaoming Yang, Li Yu and Liping Qu produced and edited the chart. All authors contributed to data analysis, drafting or revising the article, have agreed on the journal to which the article will be submitted, gave final approval of the version to be published, and agree to be accountable for all aspects of the work.

